# Causal Associations of Metformin with Five Interleukin Receptors: Uncovering Potential Anti-inflammatory Mechanisms through Mendelian Randomization

**DOI:** 10.1101/2025.04.28.25326557

**Authors:** Wang Xiaoyong, Xia Xin, Ma Ningning, Chen Wenhui, Huang Yingjie, Wu Yujian

## Abstract

**Objective:** This study aims to investigate the causal relationships between metformin (Met) and interleukin (IL) receptors using Mendelian randomization (MR) methods, providing new theoretical evidence for the clinical application of Met.

**Methods:** Instrumental variables (IVs) were obtained from genome-wide association study (GWAS) databases. The TwoSampleMR package in R was used to perform MR analysis with the inverse variance weighting (IVW) method. Sensitivity analyses were conducted using the leave-one-out (LOO) method. The Cochran Q test was used to assess heterogeneity, and MR-Egger regression intercept and P values were used to test and correct for horizontal pleiotropy. Publication bias was assessed using funnel plots. Metformin was used as the exposure factor, and two study cohorts were assigned: a training cohort (ID: ukb-b-1 4609) and a testing cohort (ID: ukb-a-159). The outcomes were five IL receptors: IL-6 receptor subunit alpha (IL-6 sRa), IL-2 receptor subunit alpha (IL-2 sRa), IL-17 receptor B (IL-17BR), IL-12 receptor subunit beta-2 (IL-12RB2), and IL-1 receptor-like 1 (IL-1RL1).

**Results:** In the training cohort analysis, all five IL receptors showed significant negative causal relationships with Met (P < 0.05). In the testing cohort analysis, Met had significant negative causal relationships with IL-6 sRa, IL-17BR, IL-12RB2, and IL-1RL1 (P < 0.05). No significant pleiotropy or heterogeneity was detected, and the results were robust.

**Conclusion:** This study reveals significant negative correlations between Met and the five IL receptors through MR analysis, suggesting that Met may directly or indirectly modulate IL receptors to exert its potential therapeutic effects. This provides new theoretical evidence for the application of Met in inflammation-related diseases.

## Introduction

Metformin (Met), since its introduction to clinical practice in 1957, has become an essential medication in the treatment of type 2 diabetes [1]. Major diabetes guidelines worldwide recommend that, unless contraindicated, patients should initiate treatment with Met and include it in all combination therapy regi mens [2, 3]. Met is not only the first-line treatment for overweight or obese patients with type 2 diabetes but is also suitable for non-overweight/non-obese individuals with the condition, with its efficacy and adverse reactions being ind ependent of the patient’s BMI [4, 5]. In recent years, studies have identified multiple non-glycemic benefits of Met, including cardiovascular protection, anti-tumor effects, improvement of polycystic ovary syndrome, weight reduction, and reduced risk of dementia [6, 7]. Additionally, Met offers favorable health economic benefits, with better cost-effectiveness [8].

In recent years, the anti-inflammatory properties of Met have become a research focus, particularly in chronic inflammatory pathways mediated by interleukin (IL), where it shows potential for intervention. IL plays a significant role in the occurrence, development, and progression of numerous non-infectious chronic diseases. Recent studies [9-11] have found that IL is closely related to cardiovascular and cerebrovascular diseases, neurological diseases, digestive system diseases, kidney diseases, autoimmune diseases, and even aging, tumor development, and metabolic disorders. For example, in cardiovascular and cerebrovascular diseases [12], chronic inflammation can promote the formation and instability of atherosclerotic plaques, thereby increasing the risk of thrombosis. In neurodegenerative diseases such as Alzheimer’s disease [13], neuroinflammation is one of the key factors driving disease progression. Moreover, inflammation is closely linked to various metabolic diseases [14], such as obesity, type 2 diabetes, and non-alcoholic fatty liver disease. The chronic low-grade inflammatory state in these diseases can further exacerbate insulin resistance and metabolic disorders.

Interleukin receptors play a crucial role in regulating inflammatory responses and are widely distributed in various cell types, including immune cells, endothelial cells, and parenchymal cells. Abnormal activation or dysfunction of IL receptors is closely associated with the occurrence and development of many inflammation-related diseases [15, 16]. Although Met has been proven to regulate inflammatory responses through various mechanisms [7, 17], research on its relationship with IL receptors remains limited [18]. In-depth studies on the interaction between Met and IL receptors will not only help reveal its potential anti-inflammatory mechanisms but also provide important evidence for developing new anti-inflammatory treatment strategies. Therefore, we employed Mendelian randomization (MR) methods to further assess the causal relationship between Met and IL receptors. This is of great significance for optimizing its clinical application in inflammation-related diseases, identifying new therapeutic targets, and improving the theoretical framework of anti-inflammatory mechanisms.

Mendelian randomization, as an innovative epidemiological research method, uses genetic variations as instrumental variables to effectively avoid confounding biases and reverse causality issues. Based on the principle of random allocation of genetic variants, this method simulates the conditions of a randomized controlled trial, providing strong evidence for causal inference. This study aims to explore the causal relationship between Met and IL receptors using MR methods to provide new theoretical evidence for the clinical application of Met and further explore its potential therapeutic effects in inflammation-related diseases.

## 1. Methods and Data

### 1.1 Study Design

This study utilized Mendelian randomization for secondary data analysis of existing databases. MR is a method that uses genetic variations of exposure as instrumental variables (IVs) to identify causal relationships between exposure phenotypes and outcomes. It leverages accessible public datasets from large-scale genome-wide association studies (GWAS) for “exposure” and “outcome” to compensate for the shortcomings of observational studies. MR analysis is designed based on the following three core assumptions:

(1) Relevance Assumption: Genetic variants (single nucleotide polymorphisms, SNPs) must have a strong association with the exposure factor to ensure that the selected SNPs can serve as effective proxy variables for the exposure [19]. To screen for SNPs strongly associated with the exposure, this study used a significance threshold of p < 5×10^-8. Subsequent analyses further validated the robustness of the results through sensitivity analysis and heterogeneity testing. F statistics were calculated to estimate sample overlap effects and weak instrument bias, requiring F > 10 [20].

(2) Independence Assumption: IVs must be independent of any known or unknown confounding factors to avoid interference from confounders in causal associations and ensure accurate causal inferences [21]. During the data screening phase, we followed the independence principle in MR analysis, ensuring that exposure and outcome data came from independent samples to avoid sample overlap bias. In the data preprocessing phase, we removed palindromic SNPs with intermediate allele frequencies. These SNPs, which have the same base arrangement in both forward and reverse sequences (e.g., A/T and G/C), may intro duce uncertainty in analysis results due to sequence structure. Additionally, we set a linkage disequilibrium (LD) threshold of r^2^ = 0.001 to ensure the independence of instrumental variables, meaning that SNPs with an r^2^ value greater than 0.001 would be considered highly correlated, and only one would be retained in the same cluster [22]. The physical distance threshold was set at kb=10M b, meaning that within a 10Mb genomic distance, the SNP with the smallest P -value would be retained, while others would be considered highly correlated a nd removed.

(3) Exclusivity Assumption: Ensuring that SNPs only affect the outcome through the exposure factor and not through other direct causal pathways. This study comprehensively validated the validity of the exclusivity assumption through horizontal pleiotropy detection, heterogeneity assessment, and sensitivity analysis, thereby ensuring the accuracy of causal inferences and the robustness of results between Met and five inflammatory factor receptors [23].

These steps collectively ensure that IVs are strongly associated with the exposure factor and are independent of each other, thereby enhancing the accuracy and reliability of Mendelian randomization studies [24].

### 1.2 Statistical Analysis Methods

The two-sample MR analysis was conducted using R software (version 4.4. 2) and the TwoSampleMR package (version 0.6.8). In this study, the Inverse Variance Weighting Fixed Effects (IVW-FE) method was selected for MR analysis. Additionally, several other methods were used to assist in estimating causal effects, including Weighted median (WMed), MR-Egger regression (MER), Simple model (SM), Weighted model (WM), and Inverse Variance Weighting Random Effects (IVW-RE). The consistency of effect sizes (β values) from WMed, MER, SM, WM, and IVW-RE with IVW indicates robust results.

In this study, we used the mr() function from the TwoSampleMR package for Mendelian randomization analysis. To ensure statistical power, we only selected two samples containing more than 10 eligible SNPs. For each eligible two-sample, we calculated the log odds ratio (LogOR) and its 95% confidence interval (CI) to assess the causal association between the exposure factor and the outcome. We also calculated the corresponding P-value for LogOR through the statistical testing module of the mr() function to assess the significance of the causal association. A P-value < 0.05 was considered statistically significant, indicating a possible causal association between the exposure factor and the out come. The magnitude of LogOR and 95% CI indicates the strength of the association, with positive LogOR values indicating a positive correlation between the exposure factor and the outcome, and negative values indicating a negative correlation.

We used the Cochran Q test to assess heterogeneity, with a P-value > 0.0 for the Q statistic indicating no significant heterogeneity [23]. Additionally, we used the intercept and P-value from MR-Egger regression to test and correct for horizontal pleiotropy, with P > 0.05 suggesting no significant horizontal pleiotropy. For two samples with significant heterogeneity or horizontal pleiotropy, we excluded them to reduce bias, enhance result robustness, and meet the basic assumptions of Mendelian randomization. Moreover, we conducted sensitivity analysis using the Leave-One-Out (LOO) method to evaluate result stability by excluding one instrumental variable at a time.

### 1.2 Data Sources

The data for this study were sourced from the IEU OpenGWAS project (h ttps://gwas.mrcieu.ac.uk/), which provides a wealth of GWAS summary data for MR analysis. We searched for GWAS datasets related to Met and IL receptor s based on the trait items in the GWAS database, identifying two datasets related to Met (GWAS IDs: ukb-b-14609, ukb-a-159) and 104 datasets related to IL receptors (see Appendix 1). To ensure the robustness and reliability of the results, we assigned Met as the exposure factor and designated a training cohort (GWAS ID: ukb-b-14609) and a test cohort (GWAS ID: ukb-a-159). Using the training cohort dataset and IL receptor-related datasets for MR analysis, we ulti mately selected five IL receptors: IL-6 receptor subunit alpha (Interleukin-6 receptor subunit alpha, IL-6 sRa), IL-2 receptor subunit alpha (Interleukin-2 receptor subunit alpha, IL-2 sRa), IL-17 receptor B (interleukin-17 receptor B, IL-17 BR), IL-12 receptor subunit beta-2 (Interleukin-12 receptor subunit beta-2, IL-1 2RB2), and IL-1 receptor-like 1 (Interleukin-1 receptor-like 1, IL-1RL1). We an alyzed the relevant outcome datasets (see Table 1) and further validated the ass ociations between these five outcome datasets and Met using the test cohort da taset.

**Table 1.** GWAS Trait and GWAS ID are the trait items and corresponding IDs from the IEU OpenGWAS project database. PMID is the unique digital identifier from the PubMed database (https://pubmed.ncbi.nlm.nih.gov). Entr ez is the unique identifier for the gene in the Entrez Gene database (https://www.ncbi.nlm.nih.gov/gene). UniProt is the unique identifier for the protein in the UniProt database (https://www.uniprot.org/uniprotkb).

| GWAS trait | GWAS ID | population | year | SNPs | PMID | Entrez | uniprot | Gene Name |
| --- | --- | --- | --- | --- | --- | --- | --- | --- |
| IL-6 sRa | prot-c-4139_71_2 | Europe | 2019 | 501 428 | 28 240 26<br>9 | 3570 | P08887 | IL6R |
| IL-2 sRa | prot-c-3151_6_1 | Europe | 2019 | 501 428 | 28 240 26<br>9 | 3559 | P01589 | IL2RA |
| IL-17B R | prot-c-5084_154_3 | Europe | 2019 | 501 428 | 28 240 26<br>9 | 55540 | Q9NRM6 | IL17RB |
| Interleukin-12 receptor subunit beta-2(IL-12RB2) | prot-a-1474 | Europe | 2018 | 10 534 735 | 29 875 48<br>8 | 3595 | Q99665 | IL12RB2 |
| Interleukin-1 receptor-like 1(IL-1RL1) | prot-a-1502 | Europe | 2018 | 10 534 735 | 29 875 48<br>8 | 9173 | Q01638 | IL1RL1 |

#### 1.2.1 Exposure Data Sets

Training cohort (GWAS ID: ukb-b-14609): This dataset, derived from a 2018 study by Ben Elsworth et al., covers 462,933 individuals of European descent, including 11,552 Met users and 451,381 non-users, analyzing 9,851,867 SNPs.

Test cohort (GWAS ID: ukb-a-159): This dataset, from a 2017 study by the Neale Lab, encompasses 337,159 individuals of European descent, with 8, 392 Met users and 328,767 non-users, analyzing 10,894,596 SNPs.

#### 1.2.2 Outcome Data Sets

The selected IL receptors and their corresponding dataset information are shown in the table below:

## 2 Results

### 2.1 Training Cohort: Significant Negative Causal Associations Between Met and Five IL Receptors

In the training cohort (exposure factor GWAS ID: ukb-b-14609), the F-values calculated for the association between SNPs and Met ranged from 30.25 to 5 77.91. The IVW-FE analysis indicated significant negative causal relationships between Met and all five IL receptors (P < 0.05). The Q statistic showed no evidence of significant heterogeneity (P > 0.05), and MR-Egger regression analysis did not detect pleiotropy (P > 0.05) (see Figure 2). The MR scatter plots suggested consistent directions (see Figure 3). During the LOO sensitivity analysis, the confidence intervals included 0 for some individual SNPs in the IL-17B R and IL-1RL1 groups, but the overall direction remained consistent (see Figure 4), confirming the robustness of the results. The consistency of β values from WMed, MER, SM, WM, and IVW-RE with IVW-FE indicated robust results (see Figure 5). Funnel plot analysis showed no obvious asymmetry, indicating no publication bias (see Figure 6).

**Figure 1:**
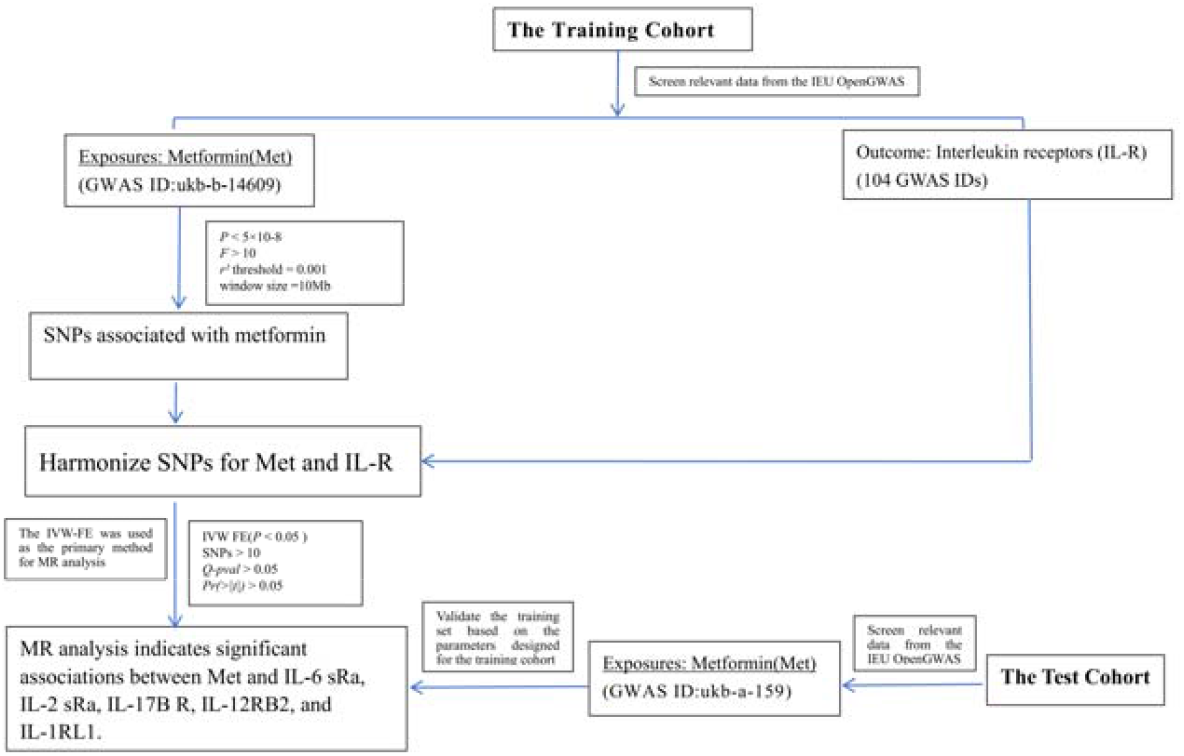
Flowchart of the study

**Figure 2:**
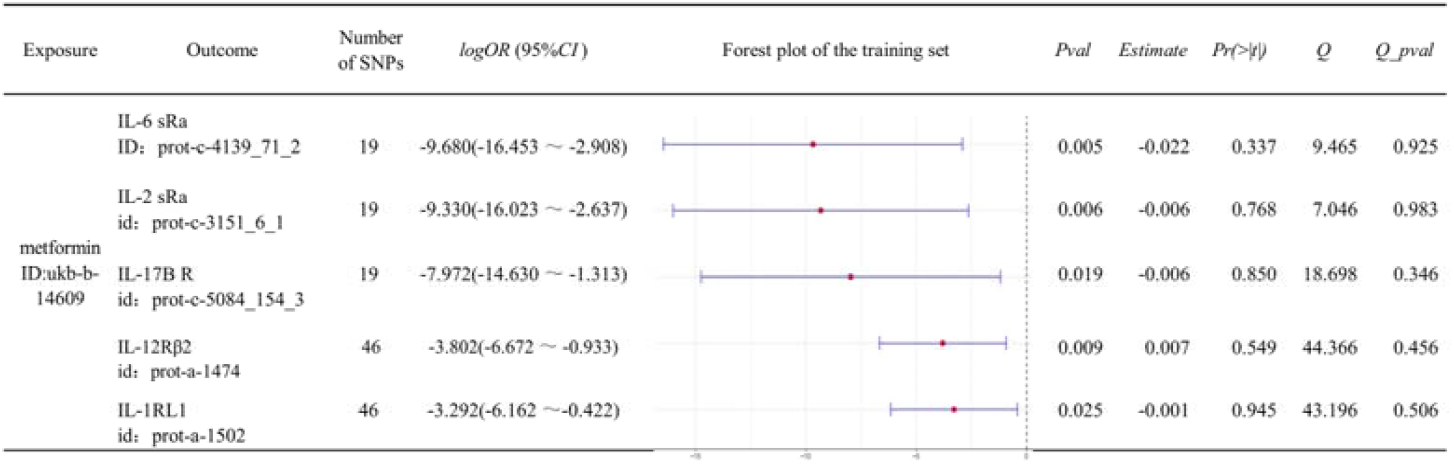
Forest plots of associations between Met and IL-6 sRa, IL-2 sR a, IL-17BR, IL-12RB2, IL-1RL1 in the training cohort, along with Q statistics (Q) and corresponding P-values (Q-pval), MR-Egger regression intercepts (Estimate) and corresponding P-values (Pr(>|t|)).

**Figure 3:**
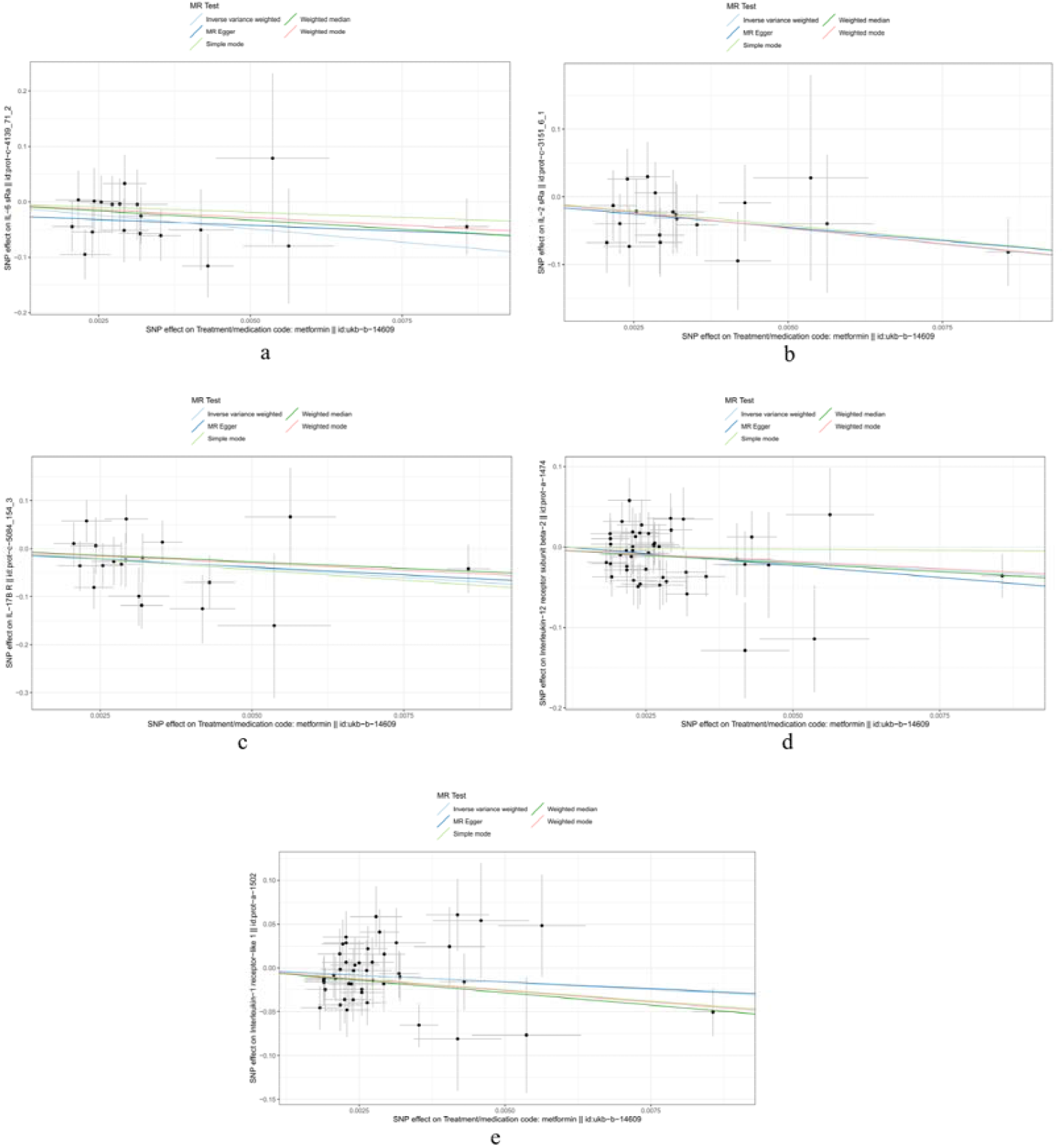
MR scatter plots of associations between Met and four IL receptors (a: IL-6 sRa, b: IL-2 sRa, c: IL-17BR, d: IL-12RB2, e: IL-1RL1) in the training cohort.

**Figure 4:**
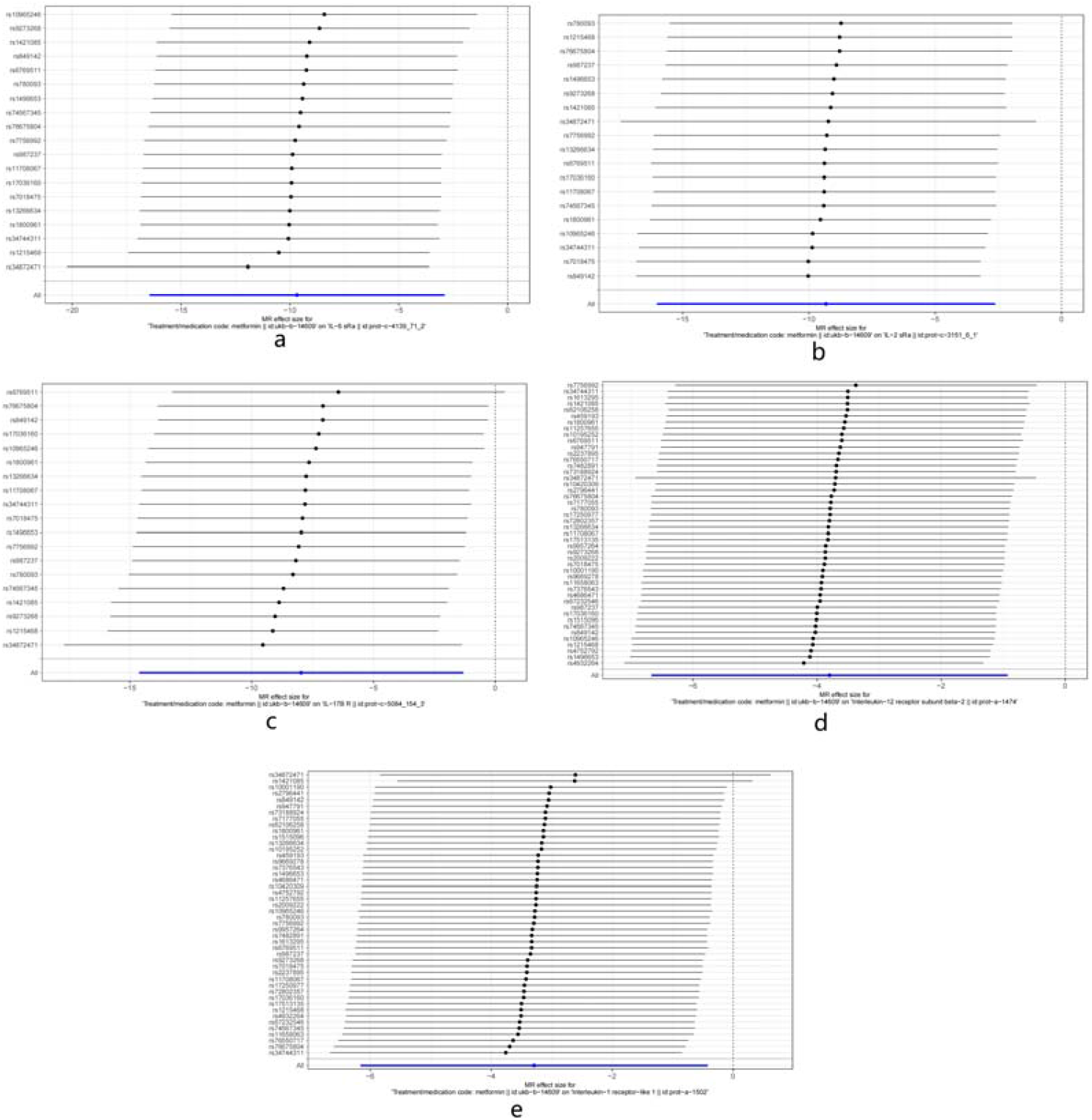
LOO sensitivity analysis forest plots of associations between Met and four IL receptors (a: IL-6 sRa, b: IL-2 sRa, c: IL-17BR, d: IL-12RB2, e: IL-1RL1) in the training cohort.

**Figure 5:**
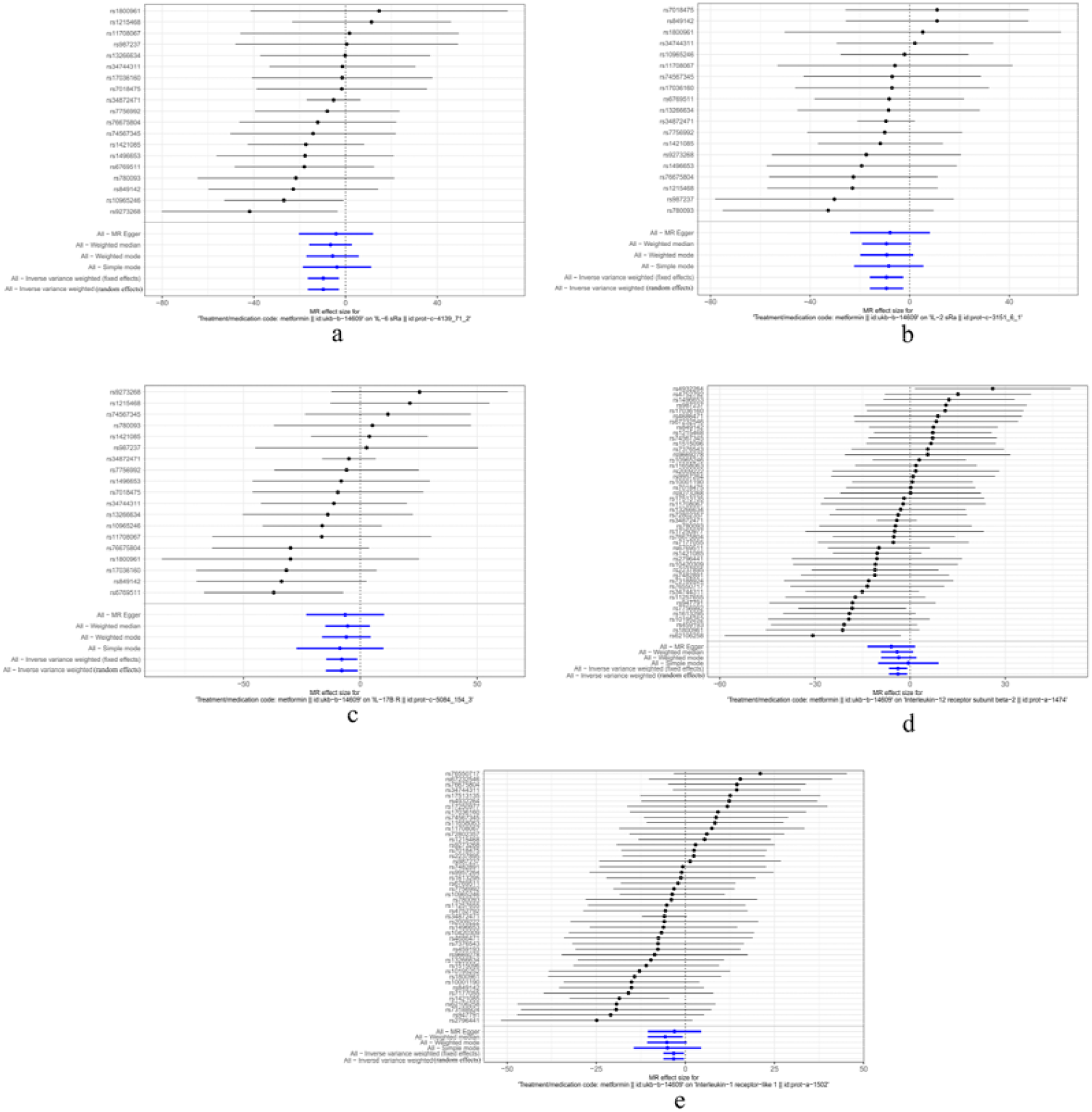
Forest plots of focal heterogeneity scores (HS) for each addition al SNP and β value directionality from MER, WMed, WM, SM, IVW-FE, IV W-RE analysis methods (a: IL-6 sRa, b: IL-2 sRa, c: IL-17BR, d: IL-12RB2, e: IL-1RL1) in the training cohort.

**Figure 6:**
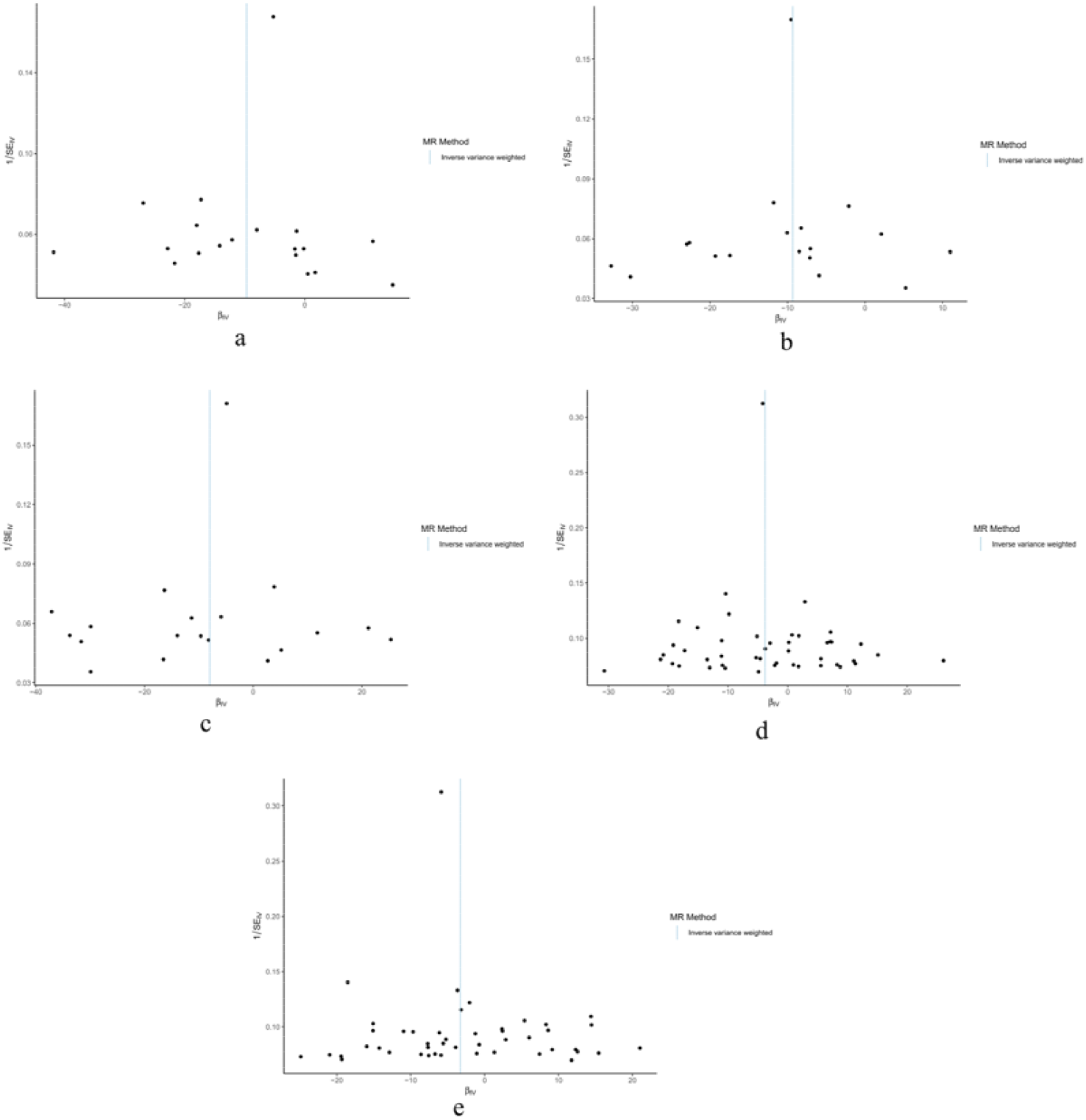
Funnel plots showing no asymmetry (a: IL-6 sRa, b: IL-2 sRa, c: IL-17BR, d: IL-12RB2, e: IL-1RL1) in the training cohort.

### 2.2 Testing Cohort: Causal Associations Between Met and IL Receptors

In the testing cohort (exposure factor GWAS ID: ukb-a-159), the F-values calculated for the association between SNPs and Met ranged from 30.81 to 45 9.26. The IVW-FE analysis indicated significant negative causal relationships between Met and four IL receptors (IL-6 sRa, IL-17BR, IL-12RB2, IL-1RL1) (P < 0.05). The Q statistic showed no evidence of significant heterogeneity, and MR-Egger regression analysis did not detect pleiotropy (see Figure 7). The MR scatter plots suggested consistent directions (see Figure 8). During the LOO sensitivity analysis, the confidence intervals included 0 for some individual SNPs in the IL-17BR and IL-1RL1 groups, but the overall direction remained consistent (see Figure 9), confirming the robustness of the results. The consistency of β values from WMed, MER, SM, WM, and IVW-RE with IVW-FE indicated robust results (see Figure 10). Funnel plot analysis showed no obvious asymmetry, indicating no publication bias (see Figure 11). However, no significant causal association was found between Met and IL-2 sRa (logOR = -5.067, 95% CI: -12.297 to 2.162, P = 0.170).

**Figure 7:**
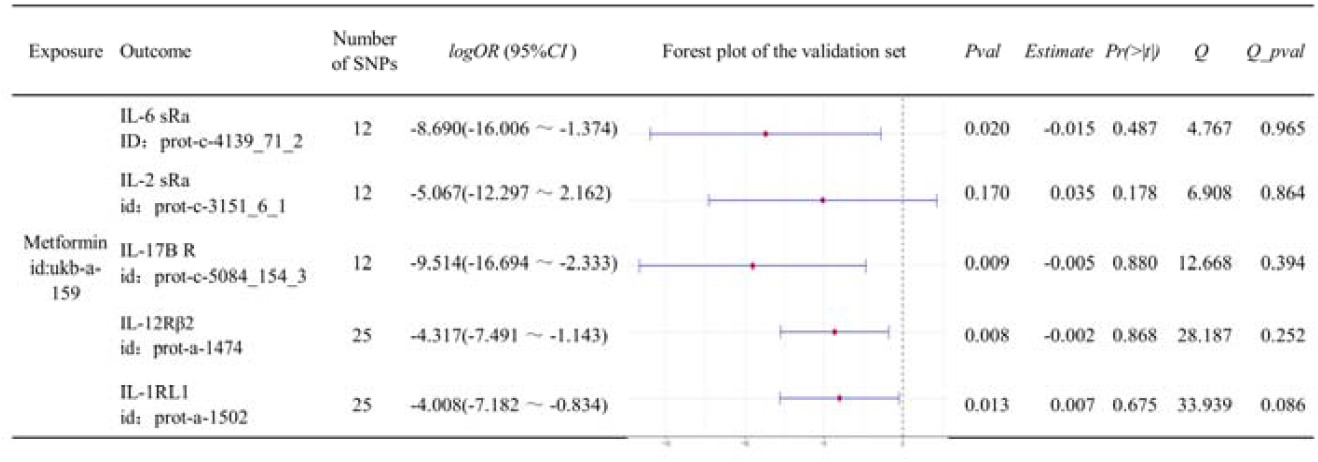
Forest plots of associations between Met and IL-6 sRa, IL-2 sR a, IL-17BR, IL-12RB2, IL-1RL1 in the testing cohort, along with Q statistics (Q) and corresponding P-values (Q-pval), MR-Egger regression intercepts (Estimate) and corresponding P-values (Pr(>|t|)).

**Figure 8:**
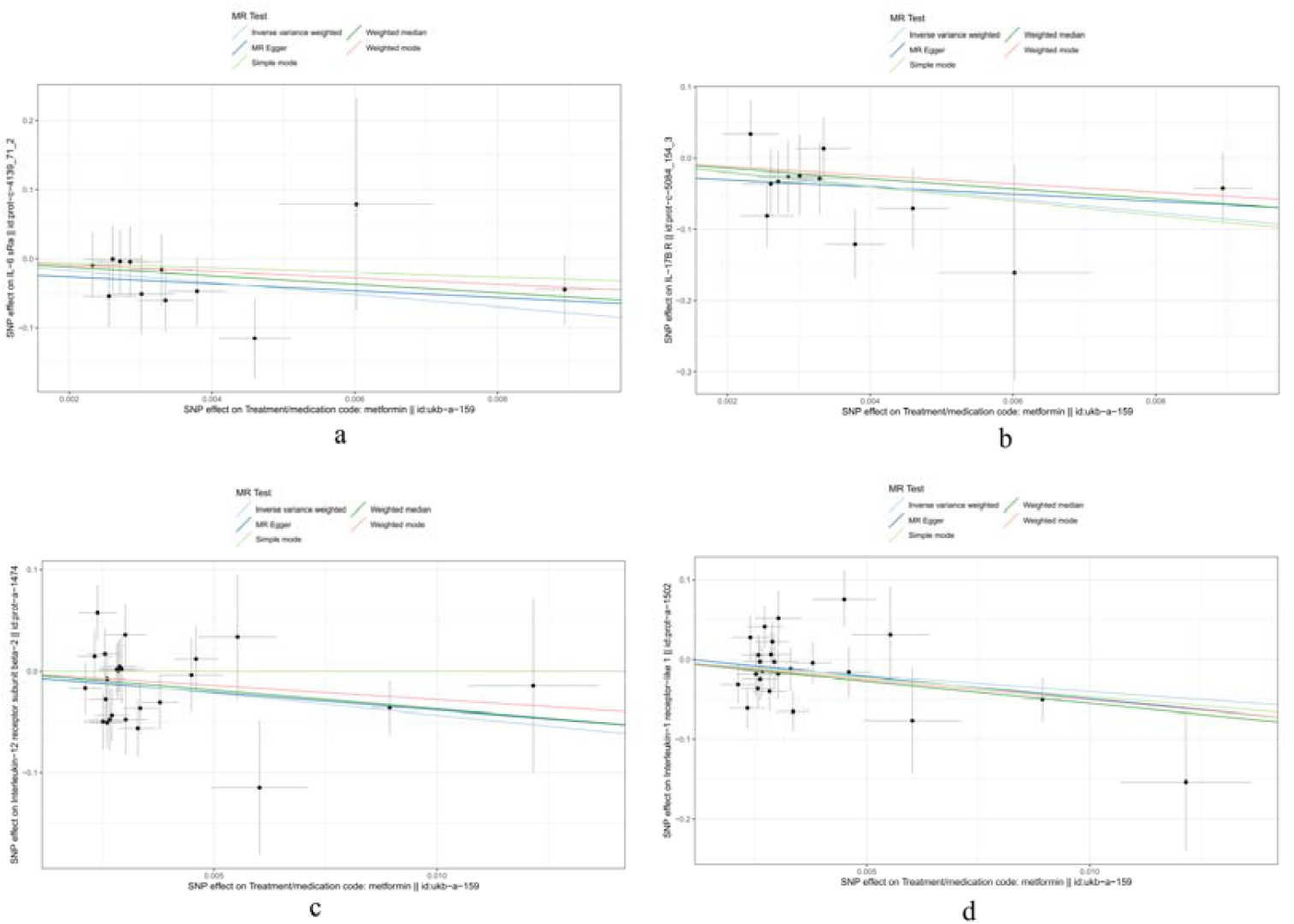
MR scatter plots of associations between Met and four IL recept ors (a: IL-6 sRa, b: IL-17BR, c: IL-12RB2, d: IL-1RL1) in the testing cohort.

**Figure 9:**
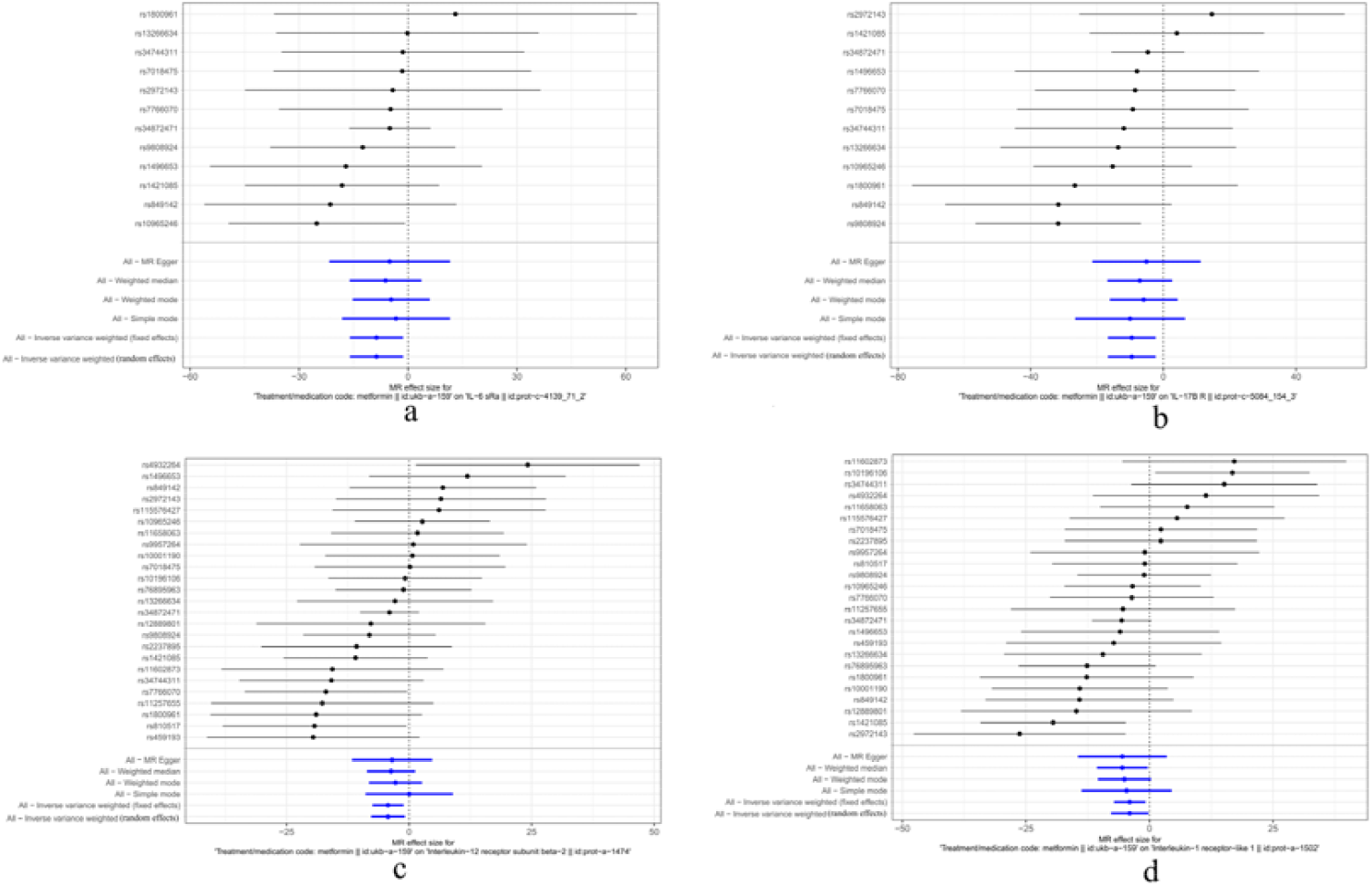
LOO sensitivity analysis forest plots of associations between Met and four IL receptors (a: IL-6 sRa, b: IL-17BR, c: IL-12RB2, d: IL-1RL1) in the testing cohort.

**Figure 10:**
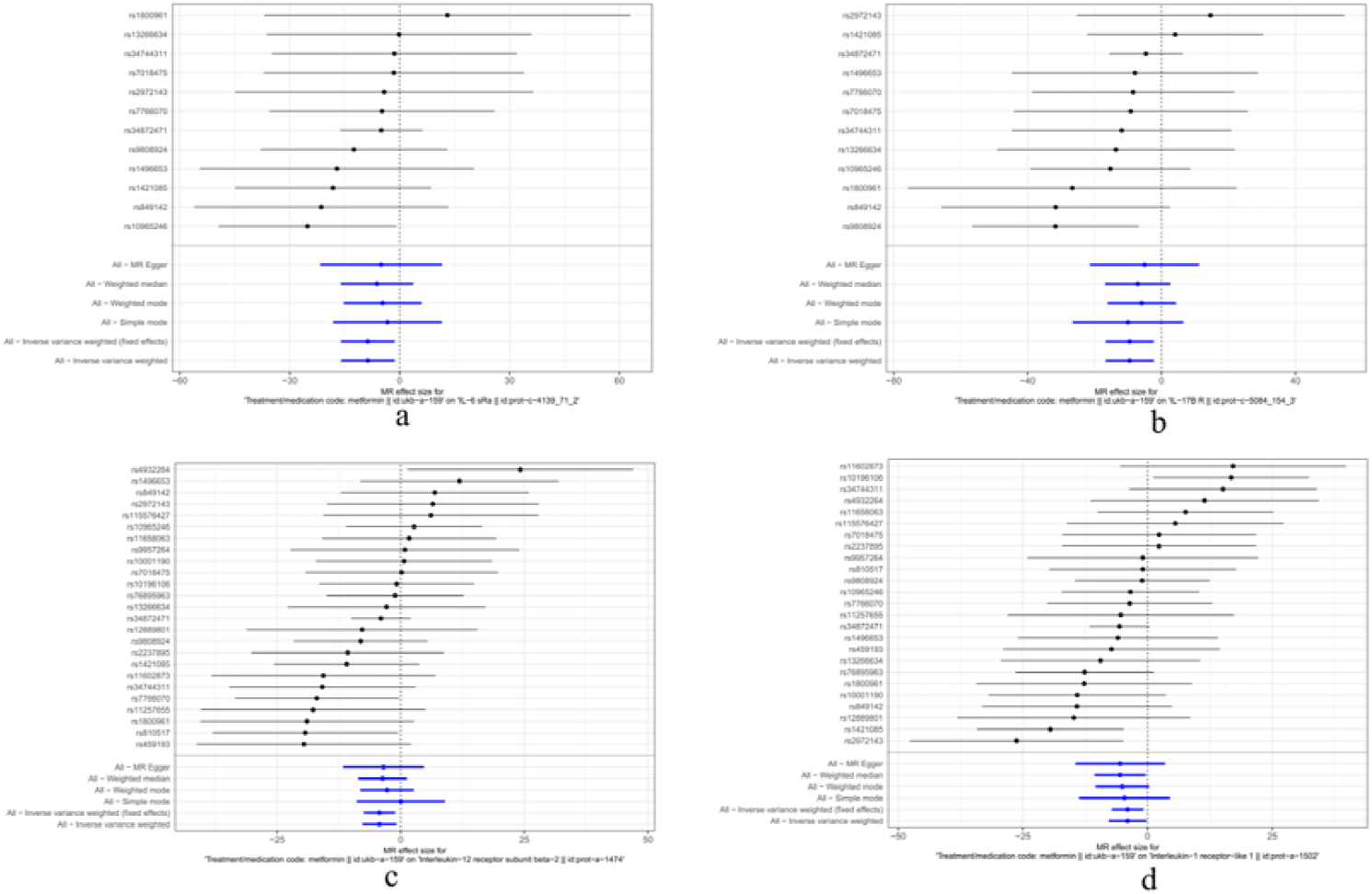
Forest plots of focal heterogeneity scores (HS) for each additional SNP and β value directionality from MER, WMed, WM, SM, IVW-FE, IV W-RE analysis methods (a: IL-6 sRa, b: IL-17BR, c: IL-12RB2, d: IL-1RL1) i n the testing cohort.

**Figure 11:**
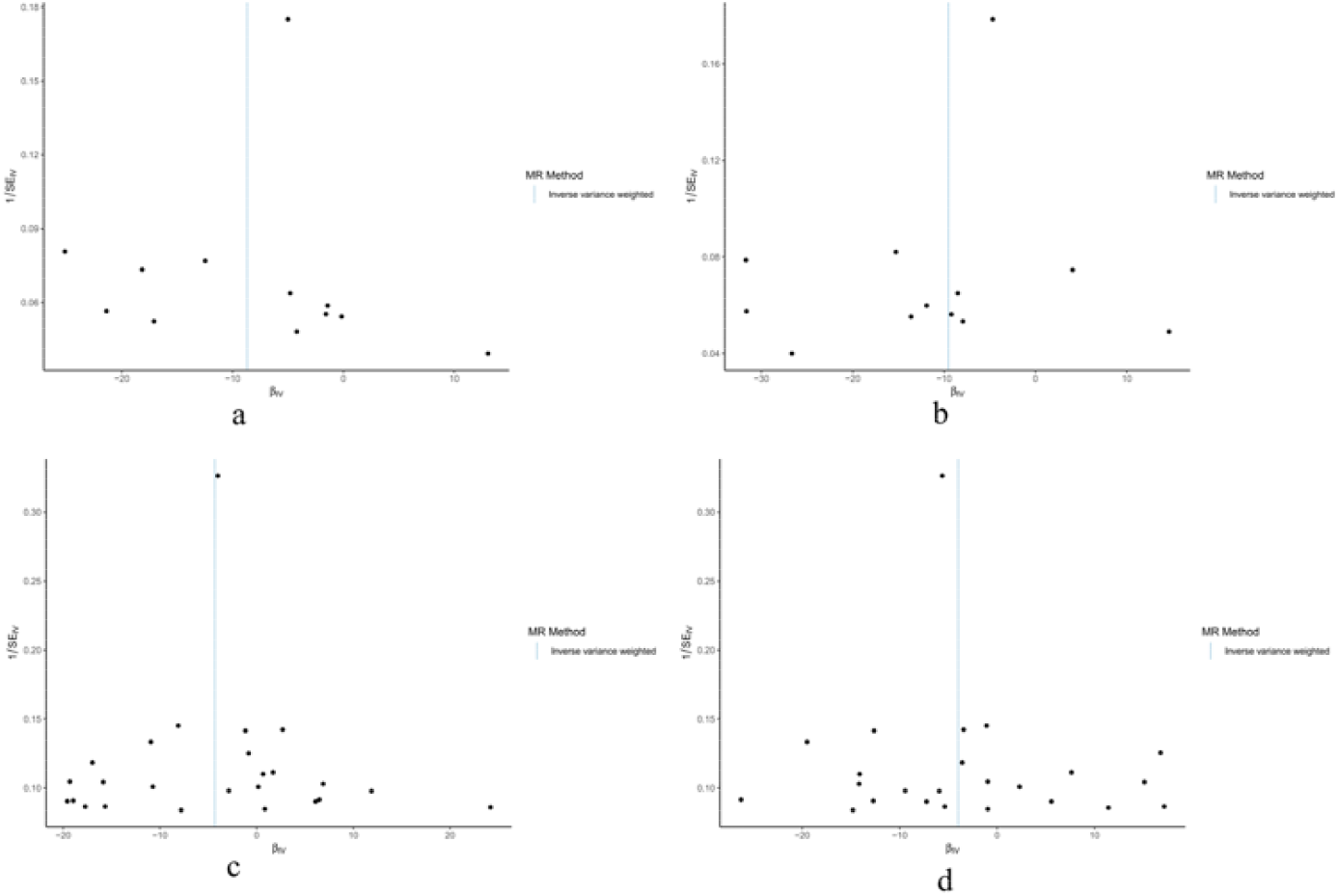
Funnel plots showing no asymmetry (a: IL-6 sRa, b: IL-17BR, c: IL-12RB2, d: IL-1RL1) in the testing cohort.

## 3 Discussion

Metformin exerts anti-inflammatory effects through multiple mechanisms, including activating the AMPK signaling pathway, regulating immune cell functions, modulating cytokines and chemokines, regulating mitochondrial function, modulating gut microbiota, inhibiting the NLRP3 inflammasome, promoting autophagy, regulating apoptosis, and modulating inflammation-related signaling pathways [7, 17]. These mechanisms collectively reduce inflammatory responses and improve symptoms of various inflammatory diseases. This study, for the first time, used Mendelian randomization to assess the causal relationships between Met and five interleukin receptors (IL-6sRα, IL-2sRa, IL-17BR, IL-12RB2, IL-1 RL1), revealing significant negative causal relationships. This suggests that Met may modulate the expression or activity of these inflammatory receptors to exert anti-inflammatory effects, providing new theoretical evidence for the potential application of Met in anti-inflammatory and diabetes complication management.

### 3.1 Negative Causal Relationship Between Met and IL-6sRα and Its M echanisms

IL6Rα (also known as CD126) is a component of the IL-6 receptor, existing in both membrane-bound (mIL-6R) and soluble forms (sIL-6R). IL-6 mediates signaling pathways via membrane-bound and soluble receptors, playing roles in immune regulation, hematopoiesis, inflammation, tumor development, metabolic control, and sleep [25, 26]. In the classical signaling mode, IL-6 binds to mIL-6R and recruits IL-6 signal transducer (IL6ST) to form a complex, activating JAK-STAT and Ras-MAPK signaling pathways to regulate cell proliferation, differentiation, and acute-phase responses [27]. In trans-signaling, sIL-6R complexes with IL-6 can directly act on cells expressing IL6ST, significantly expanding the signaling range and primarily participating in pro-inflammatory responses [28].

The negative causal relationship between Met and IL-6sRα suggests that Met may inhibit the expression or function of mIL-6R or sIL-6R, thereby bloc king IL-6-mediated inflammatory signaling. This is consistent with previous findings by Mishra AK et al. [29], who reported that Met specifically reduces IL-6R expression through AMPK, mTOR, and miR-34a. The specific mechanisms may include: (1) AMPK-dependent inhibition of mTORC1 activation, blocking the JAK/STAT3 signaling pathway; (2) AMPK-independent inhibition of mTORC1 activation, potentially regulating the Ras-MAPK signaling pathway; and (3) reducing IFN-α-mediated apoptosis by inhibiting the Ras/ERK signaling pathway, indirectly regulating JAK/STAT3 activity. These findings indicate that Met may modulate the IL-6 signaling pathway through multiple targets to exert anti -inflammatory and anti-tumor effects.

### 3.2 Negative Causal Relationship Between Met and IL-2sRa and Its Mechanisms

IL-2sRa (also known as CD25 or p55) is a key subunit of the IL-2 receptor complex. It activates the JAK-STAT signaling pathway (especially STAT5) upon binding to IL-2, regulating T cell activation, proliferation, and differentiation, and participating in the pathogenesis of autoimmune diseases, tumor immune evasion, and inflammatory responses [30].

The significant negative causal relationship between Met and IL-2sRa, revealed by MR analysis, provides a new molecular basis for Met’s immune regulatory effects. Studies [31, 32] suggest that Met may regulate IL-2 effects by influencing the activity of the JAK-STAT signaling pathway, potentially involving direct or indirect inhibition of JAK kinase activity: (1) Met may directly or in directly inhibit JAK kinase activity, blocking STAT5 phosphorylation and weakening IL-2 signal transmission; (2) Met may regulate IL-2sRa expression levels through epigenetic modifications or transcription factor control, reducing the binding efficiency of IL-2 to its receptor. These mechanisms collectively inhibit the overactivation of T cells, reduce the secretion of pro-inflammatory cytokines, and potentially enhance the immunosuppressive functions of regulatory T cells (Tregs).

From a biological perspective, Met’s modulation of the IL-2 signaling path way has clear clinical significance. By inhibiting the differentiation of Th1 and Th17 cells, Met may alleviate immune pathological damage in autoimmune diseases such as rheumatoid arthritis and inflammatory bowel disease [32]. Additionally, by maintaining Treg stability, Met may enhance immune tolerance and prevent immune-related adverse reactions. Met’s anti-inflammatory effects may also extend to the tumor microenvironment, where it may interfere with tumor cell immune evasion mechanisms by inhibiting T cell-mediated inflammatory responses.

### 3.3 Negative Causal Relationship Between Met and IL-17BR and Its Mechanisms

IL-17BR, a crucial member of the IL-17 receptor family, specifically binds IL-17B and IL-17E and is widely expressed in various endocrine tissues. This receptor activates downstream JAK-STAT and MAPK signaling pathways and triggers the NF-kappaB signaling pathway to promote IL-8 production, thereby exerting multiple biological effects, including anti-inflammatory actions, tumor promotion, host defense, and asthma development [33]. IL-17BR plays a key role in the pathogenesis of various autoimmune diseases (e.g., rheumatoid arthritis, psoriasis), where its abnormal activation or expression is often closely related to disease progression [34].

The significant negative causal relationship between Met and IL-17BR indi cates that Met may inhibit the binding of IL-17 to its receptors (including IL-1 7BR), thereby reducing inflammatory responses. This association is supported by several studies [35, 36], suggesting that Met can influence the IL-17 signaling pathway to modulate immune responses. Specific mechanisms may include: (1) Met’s activation of SIRT1 in cancer treatment [37], potentially reducing Th17 cell frequency and thereby affecting IL-17 signaling pathway activity; (2) Met’s ability to lower inflammatory parameters, increase FoxP3 and TGF-β protein levels, and reduce IL-17 expression, indicating that it may promote Treg cell differentiation or function, regulate the Th17/Treg cell balance, and inhibit excessive inflammatory responses [38].

### 3.4 Negative Causal Relationship Between Met and IL-12RB2 and Its Mechanisms

IL-12Rβ2, as a core component of the IL-12 receptor, forms a high-affinity binding site with IL-12Rβ1 to mediate IL-12-dependent signaling. IL-12 plays a crucial role in the differentiation of naive T cells into Th1 cells. IL-12 enhances the cytotoxicity of NK cells and CD8+ T cells and promotes IFN-γ-dependent CXCL10 generation, exerting anti-angiogenic activity. IL-12Rβ2 expression is bidirectionally regulated by Th1/Th2 differentiation-related cytokines, and its activation promotes Th1 differentiation and IFN-γ production through the JA K-STAT pathway, while also participating in immune regulation as part of the IL-35 receptor complex [39].

He W et al. [40] reported that Met downregulates CD8+ T cells to inhibit hepatocellular carcinoma in non-diabetic individuals; Petrovic AR et al. [41] reported that Met directly enhances NK cell activation and cytotoxicity. Although no studies have reported the association between Met and IL-12RB2, MR analysis in this study indicates a significant negative causal relationship between Met and IL-12RB2. However, the mechanisms are unclear. Met may modulate immune responses through the following IL-12RB2-related mechanisms: (1) Inhibiting Th1 cell differentiation by reducing IL-12Rβ2 expression or activity, thereby decreasing IFN-γ production and suppressing Th1-mediated inflammation; (2) Affecting tumor angiogenesis by blocking IL-12’s induction of CXCL10 through IFN-γ; (3) Regulating NK/CD8+ T cell cytotoxicity by interfering with I L-12Rβ2-mediated signaling, impacting tumor immune surveillance; (4) Modulating IL-35 signaling pathways by influencing Treg differentiation and function through IL-12Rβ2.

The negative causal association between Metformin and IL-12RB2 reveals its potential role in immune regulation, offering a new perspective for developing novel therapeutic strategies. However, further research is needed to elucidate how Metformin specifically affects IL-12RB2 function and how these effects translate into clinical therapeutic outcomes.

### 3.5 Negative Causal Relationship Between Met and IL-1RL1 and Its Mechanisms

IL-1RL1 (also known as ST2) is a member of the IL-1 receptor family, existing in both membrane-bound (ST2L) and soluble (sST2) forms. Upon activation by its ligand IL-33, IL-1RL1 can modulate the PI3K/AKT signaling pathway, thereby influencing cell survival and function. IL-33, as the specific ligand for ST2, primarily activates Th2-type immune responses, promoting the secretion of IL-5, IL-13, and other cytokines by Th2 cells while regulating the activity of various immune cells [42]. IL-1RL1 is widely involved in the pathogenesis of multiple chronic inflammatory diseases, including allergic diseases, autoimmune diseases, inflammatory bowel diseases, cardiovascular diseases, liver diseases, malignant tumors, and obesity-related metabolic diseases [43].

The negative causal relationship between Met and IL-1RL1 suggests that Met may modulate IL-1RL1 activity to inhibit inflammatory signaling. Zhang L et al. [44] reported that Met reduces IL-33 levels in patients with diabetic nephropathy, and Asensio-Lopez MC et al. [45] reported that Met lowers sST2 levels. Although no studies have reported the mechanisms of these associations, given the known mechanisms of IL-1RL1/IL-33 and the pharmacological actions of Met, we speculate that the association mechanisms may involve: Met activating AMPK to indirectly influence the PI3K/AKT signaling pathway, and Met’s immune regulatory effects impacting IL-33 signaling pathways involved in immune responses and inflammatory processes, thereby participating in various physiological and pathological processes.

### 3.6 Limitations and Future Research Directions

Despite the use of Mendelian randomization in this study, several limitations exist. First, the selection and quality of instrumental variables are crucial for the accuracy of the results. Although high-quality instrumental variables were selected through strict significance thresholds, clumping processes, and F statistics, weak instrumental variables or horizontal pleiotropy may still affect the robustness of the results. Second, the study is based on GWAS data from European populations, and the generalizability of the findings may be limited. Genetic backgrounds and environmental factors vary across different races and populations, potentially influencing the causal relationships between Met and inflammatory markers. Third, in the validation cohort, no significant causal association was found between Met and IL-2sRa, which may be related to sample size, genetic variant selection, or statistical method limitations and requires further research for validation. Additionally, while this study assessed the causal relationships between Met and inflammatory markers, it did not further verify the biological mechanisms. Future research should combine experimental studies and clinical trials to explore Met’s anti-inflammatory mechanisms in detail.

## 4 Conclusion

This study used Mendelian randomization to analyze the causal relationships between Met and five IL receptors (IL-6 sRa, IL-2 sRa, IL-17BR, IL-12RB2, IL-1RL1). The results indicate significant negative causal associations, suggesting that Met may exert anti-inflammatory effects by inhibiting inflammatory signaling pathways and modulating receptor activity.

These findings not only expand the potential applications of Met but also provide scientific evidence for its use in non-diabetic inflammatory diseases and offer new ideas for developing novel anti-inflammatory therapeutic strategies. However, it is important to note that the results of this study are based on Mendelian randomization analysis, which, while providing strong evidence for causal inference, still has certain limitations. Therefore, caution should be exercised when interpreting and applying the findings of this study to avoid overinterpretation. Future research could further verify the direct effects of Met on IL receptors through randomized controlled trials and elucidate the specific molecular mechanisms of its anti-inflammatory effects through in vitro cell experiment s and animal models.

## Data Availability

All data produced in the present study are available upon reasonable request to the authors
All data produced in the present work are contained in the manuscript
All data produced are available online at:https://api.opengwas.io/profile/

